# Safety and immunogenicity of a third dose of COVID-19 protein subunit vaccine (Covovax™) after homologous and heterologous two-dose regimens

**DOI:** 10.1101/2022.09.07.22279684

**Authors:** Sitthichai Kanokudom, Jira Chansaenroj, Nungruthai Suntronwong, Suvichada Assawakosri, Ritthideach Yorsaeng, Pornjarim Nilyanimit, Ratchadawan Aeemjinda, Nongkanok Khanarat, Preeyaporn Vichaiwattana, Sirapa Klinfueng, Thanunrat Thongmee, Apirat Katanyutanon, Wichai Thanasopon, Jirawan Arayapong, Withak Withaksabut, Donchida Srimuan, Thaksaporn Thatsanatorn, Natthinee Sudhinaraset, Nasamon Wanlapakorn, Sittisak Honsawek, Yong Poovorawan

## Abstract

**Objective:** To report the safety and immunogenicity profile of a protein subunit vaccine (Covovax^™^) given as a third (booster) dose to individuals primed with different primary vaccine regimens.

**Methods:** Individuals primed with two doses of COVID-19 vaccines for at least 3 months were enrolled and assigned to five groups according to their primary vaccine regimens: CoronaVac, BBIBP-CorV, AZD1222, BNT162b2, and CoronaVac/AZD1222. Immunogenicity analysis was performed to determine binding antibodies, neutralizing activity, and the T-cell response.

**Results:** Overall, 215 individuals were enrolled and boosted with the Covovax^™^ vaccine. The reactogenicity achieved was mild-to-moderate. Most participants elicited a high level of binding and neutralizing antibody responses against wild type and omicron variants following the booster dose. The 197 participants were classified by anti-N IgG. Of these, 141/197 (71.6%) were a seronegative population, and neutralizing activity and IFN-γ release were further monitored. A booster dose could elicit neutralizing activity to wild type and omicron variants by more than 95% and 70% inhibition at 28 days, respectively. The Covovax^™^ vaccine could elicit a cell-mediated immune response.

**Conclusion:** The protein subunit vaccine (Covovax^™^) can be proposed as a booster dose after two different priming dose regimens. It has strong immunogenicity and good safety profiles.

## 1. Introduction

The omicron variant (B.1.1.529) of severe acute respiratory virus 2 (SARS-CoV-2) was first identified in November 2021 (Viana et al., 2022) and has dramatically increased the transmission of Coronavirus disease 2019 (COVID-19) globally. Consequently, several vaccines targeting the SARS-CoV-2 spike (S) protein have been developed. The COVID-19 vaccine protects against serious diseases, hospitalizations, and death. However, vaccination does not entirely prevent infection and transmission to others. Massive two-dose vaccination campaigns cannot control breakthrough infections caused by the variants (Cele et al., 2022, Kuhlmann et al., 2022). A third dose is recommended to obtain high immunity against the omicron variant and its subvariants. In Thailand, several forms of COVID-19 vaccines have been introduced, including inactivated vaccines (CoronaVac, BBIBP-CorV), viral vectored vaccines (AZD1222), and mRNA vaccines (BNT162b2), which have resulted in the population receiving two-dose (prime) series of the vaccine. Several two-primed vaccine regimens have been widely used and approved. Previous studies have shown that booster doses of homologous and heterologous regimes achieved a high level of antibody titers (Munro et al., 2021, Wanlapakorn et al., 2022a) and are associated with strong protection against the omicron variant (Guirakhoo et al., 2022). Evidence has also indicated that the booster dose improved vaccine effectiveness and reduced severe outcomes of COVID-19 (Grewal et al., 2022).

On 21 April 2022, the Quadrilateral Security Dialogue donated 200,000 of doses Covovax^™^ (Serum Institute of India (SII) Pvt Ltd, Pune, India) to Thailand. This vaccine, also known as Novavax vaccine (NVX-CoV2373/ Nuvaxovid^™^) (Novavax, Inc., Gaithersburg, MD, USA) is manufactured under the brand name Covovax^™^. Covovax^™^ is a protein subunit vaccine to prevent COVID-19.

Protein subunit vaccines have been used for decades to prevent viral infectious diseases. To date, recombinant protein subunit vaccines have emerged as candidate vaccines. This vaccine, along with the adjuvant, can directly elicit protective immune responses. The protein subunit vaccine, which derives from a more traditional technology, may appear to have lower side effects in this setting, although this remains to be seen. Moreover, protein subunit vaccines present additional benefits than the viral vector and mRNA vaccines because as they overcome the limitations due to storage conditions and is easier to ship to rural/remote areas (Bhiman et al., 2022; Fiolet et al., 2022).

The United States Food and Drug Administration (US-FDA) has recently authorized the Novavax protein subunit vaccine for primary series vaccination of recipients aged 12 years and older (US-FDA. 2022). A phase 3 trial in the United Kingdom demonstrated that two doses of Novavax showed an efficacy of 86.3% against the alpha variant and 96.4% against other variants that circulated between September and November 2021 (Heath et al., 2021). Furthermore, the United States and Mexico demonstrated 90% efficacy against symptomatic and 100% efficacy against severe COVID-19 (Dunkle et al., 2022). However, this vaccine has not been authorized to be used as a booster dose in the United States. Recently, the COV-Boosted trial in the United Kingdom investigated the booster effect of the Novavax vaccine in two-dose individuals primed with AZD1222 and BNT162b2. The results showed that a high level of anti-Spike IgG could be detected in AZD1222 and BNT162 primed individuals, and neutralizing antibody against the wild type and delta variant was also evident (Munro et al., 2021). However, information is limited on the use of the COVID-19 protein subunit vaccine as the third booster dose following vaccination with different platforms and schedules, especially in combination with a series of inactivated vaccine. This study investigated the reactogenicity and immunogenicity of the protein subunit COVID-19 vaccine Covovax^™^ (Serum Institute of India Pvt Ltd, Pune, India) as a third dose (booster dose) subsequent to a two-dose primary series COVID-19 vaccine.

## 2. Materials and methods

### 2.1. Study design and participants

The cohort study was conducted between May and July 2022 at the Center of Excellence in Clinical Virology, Chulalongkorn University, Bangkok, and 10 vaccination sites in Chonburi province. The study protocol was approved by the Institutional Review Board (IRB) of the Faculty of Medicine of Chulalongkorn University (IRB 871/64) and was performed under the principles of the Declaration of Helsinki. This trial was registered in the Thai Clinical Trials Registry (TCTR 20210910002). Written informed consent was obtained from participants prior to enrollment.

A total of 215 healthy adults 18 years and older, who had previously been immunized with any two doses of COVID-19 vaccine for at least 3 months, were enrolled. The cohorts were assigned five groups according to their primary vaccine series, including homologous and heterologous regimens. All participants with no history of COVID-19 disease were reactivated with Covovax^™^ as a booster dose.

Patients were classified into five groups based on the priming vaccine regimen received: two doses of CoronaVac (hereafter referred to SVC), two doses of BBIBP-CorV (hereafter referred to SPC), two doses of AZD1222 (hereafter referred to AZC), two doses of BNT162b2 (hereafter referred to PFC), and heterologously primed with CoronaVac/AZD1222 (hereafter referred to SAC). Blood samples were collected before vaccination (day 0, baseline) and after the booster dose (day 14 ± 7 and day 28 ± 7). The sera samples were subjected to laboratory evaluation. Twelve participants were excluded from the analysis for the following reasons: in two participants the baseline anti-RBD Ig levels were unreliable (18,877 and 15,479 U/mL) and the baseline anti-nucleocapsid IgG (anti-N IgG) was negative, while 10 participants were no longer able to continue with the project.

We selected anti- N IgG for baseline specimens and divided subjects into seropositive and seronegative anti-N IgG groups. Participants who had seropositive anti-N IgG (≥ 1.4) before and after vaccination were classified into previous infection and breakthrough infection, respectively. In contrast, individuals who had seronegative anti-N IgG were further assessed for immune responses, as described in Figure 1.

**Figure 1.**
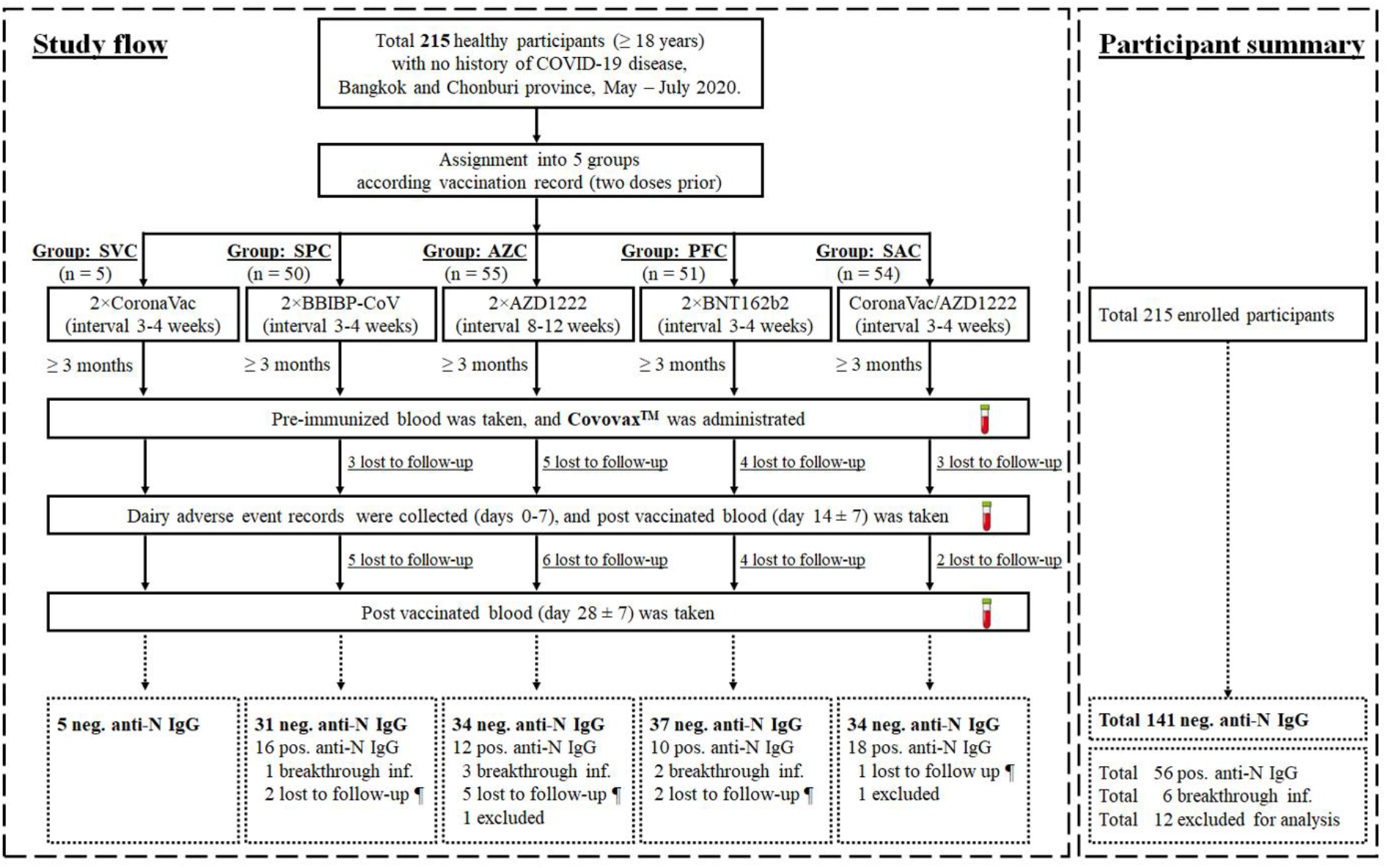
Study flow diagram of participant enrollment, blood sampling, and participant categorization. Participants who were primed with two doses of vaccines included SVC, SPC, AZC, PFC, and SAC at least 3 months after the second dose were enrolled to receive the protein subunit vaccine (Covovax^™^) as a booster dose. Blood samples were collected on days 0, 14 (±7) and 28 (±7) post booster vaccination. After observation and laboratory testing, the participants were categorized for further analysis according to the anti-N IgG titers. Only seronegative anti-N IgG participants were eligible for evaluation of immunogenicity analyses after booster vaccination. The symbol (¶) indicates participants who could not continue the study. The dotted boxes indicate the participants after categorization using anti-N IgG. neg – negative, pos – positive, inf – infection.

### 2.2. Vaccine used as a booster dose

Covovax™ was used as a booster dose in this trial. One dose (0.5 mL) contains 5 μg of SARS-CoV-2 recombinant Spike Protein (Prototype Wuhan) with 50 μg Matrix-M1™ adjuvants (Serum Institute of India, 2022).

### 2.3. Safety assessments

The participants self-reported reactogenicity using an electronic or paper questionnaire, starting on the day of vaccination and for 7 subsequent days (days 0–7). Local, systemic, and adverse events (AE) were classified as mild, moderate, and severe, as previously described (Kanokudom et al., 2022).

### 2.4. Laboratory assessments

Serum samples were collected to determine binding antibody responses, including total immunoglobulin (Ig) anti-RBD of the SARS-CoV-2 spike protein (the anti-RBD Ig), the anti-RBD IgG, and anti-N IgG as previously described (Kanokudom et al., 2022). Neutralizing activities against wild type (Euroimmun, Lubeck, Germany) and omicron (BA.2) (GenScript Biotech, NJ, USA) were analyzed using a surrogate virus neutralization test (sVNT) as previously described (Assawakosri et al., 2022b, Wanlapakorn N. et al., 2022b). The seropositivity of sVNT against wild type and omicron (BA.2) showed ≥35% and ≥30% inhibition, respectively. The interferon-gamma (IFN-γ) release assay measuring the T cell response to SARS-CoV-2 to Antigen 3 was performed using a heparinized whole blood sample. An elevated response was defined as a value of at least 0.20 IU/mL greater than the negative control (Nil), which was used to subtract IFN-γ release not deriving from SARS-CoV-2-specific T cell stimulation (Busà et al., 2022).

### 2.5. Statistical analysis

The sample size was calculated using the G* power software version 3.1.9.6 (based on conventional effect size = 0.25, given significance level (α) = 0.05, power (1-β) = 0.8, numerator degree of freedom = 3, and number of groups = 4). The calculated total sample was 179. Therefore, the desired enrolled sample with 10% dropout coverage is approximately 200 (n=50/group).

Categorical analyses of age and sex were performed using Pearson’s Chi-square test. Anti-RBD Ig and IgG were designated as geometric mean titers (GMT) with a 95% confidence interval (CI). sVNT and IFN-γ values are presented as medians with interquartile ranges (IQR). The geometric mean ratio (GMR) of anti-RBD Ig/IgG was calculated using SPC as the referent group. Significant differences between groups in antibody titers and percentage inhibition were calculated by ANCOVA with Bonferroni’s adjustment. High IFN-γ values (IU/mL minus Nil) was evaluated using the Mann–Whitney U test. A *p*-value of <0.05 was considered statistically significant.

## 3. Results

### 3.1 Demographic data and baseline characteristics

A total of 215 participants were enrolled to receive Covavax^™^ as a booster dose. According to their primary vaccine regimens, participants were classified into five groups: SVC (n=5), SPC (n=50), AZC (n=55), PFC (n=51), and SAC (n=54). The sample size of the SVC group was insufficient to conduct the statistical analysis (n=5). Hereafter, only four groups were considered for the statistical analysis. The number of women per total (%) participants was 2/5 (40.0%), 21/50 (42.0%), 29/55 (52.7%), 28/51 (54.9%), and 23/54 (42.6%), respectively. The mean age (SD) of the participants was 42.0 (11.2), 39.3 (12.3), 49.7 (14.6), 42.0 (18.7), and 37.7 (13.9), respectively. All participants presented a good healthy profile observed by the physician (well-control comorbidities were permitted) as described in Table 1. The mean (SD) of the interval between doses 2 and 3 was 367 (22.3), 236.3 (32.5), 218.9 (37.6), 185.5 (64.8), and 235.3 (31.2) for each respective group. The blood sample was collected 2 and 4 weeks after vaccination, as described in Table 1. The sex of the participants and the duration of blood collection were comparable, while the age and the interval between doses 2 and 3 showed significant differences.

**Table 1.** Demographics and baseline characteristics of enrolled participants who received Covovax^™^ as a booster dose (total 215 enrolled participants).

|  | **SVC** | **SPC** | **AZC** | **PFC** | **SAC** |
| --- | --- | --- | --- | --- | --- |
| **Total number** | 5 | 50 | 55 | 51 | 54 |
| **Sex, Female/total (%)** | 2/5 (40.0) | 21/50 (42.0) | 29/55 (52.7) | 28/51 (54.9) | 23/54 (42.6) |
| **Mean age in year (SD)** | 46.0 (11.2) | 39.3 (12.3) | 49.7 (14.6) | 42.0 (18.7) | 37.7 (13.9) |
| **No comorbidity (%)** | 3/5 (60.0) | 38/50 (76.0) | 30/55 (54.5) | 32/51 (62.7) | 45/54 (83.3) |
| **Underlying diseases (%)** |  |  |  |  |  |
| **Allergy** | - | - | 2/55 (36.4) | 1/51 (19.6) | - |
| **Asthma** | - | - | 2/55 (36.4) | - | - |
| **Cardiovascular diseases** | - | 1/50 (2.0) | 1/55 (18.2) | - | - |
| **Diabetes Mellitus** | - | 4/50 (8.0) | 11/55 (20.0) | 10/51 (19.6) | 3/54 (5.6) |
| **Dyslipidemia** | 1/5 (20.0) | 4/50 (8.0) | 6/55 (7.3) | 4/51 (7.8) | 2/54 (3.7) |
| **Hypertension** | - | 8/50 (16.0) | 13/55 (23.6) | 13/51 (25.5) | 8/54 (14.8) |
| **Thyroid diseases** | 1/5 (20.0) | - | 1/55 (18.2) | 2/51 (3.9) | - |
| **Other** | 1/5 (20.0) | 1/50 (2.0) | 3/55 (5.5) | 4/51 (7.8) | 3/54 (5.6) |
| **Interval between doses 2 and 3** |  |  |  |  |  |
| **Mean (SD)** | 367.0 (22.3) | 236.3 (32.5) | 218.9 (37.6) | 185.5 (64.8) | 235.3 (31.2) |
| **Duration of sample collection** |  |  |  |  |  |
| **(days)** |  |  |  |  |  |
| **Mean (SD) of visit 2** | 14.0 (0.7) | 14.1 (1.0) | 13.8 (1.2) | 14.0 (0.9) | 13.6 (0.9) |
| **Mean (SD) of visit 3** | 27.8 (0.5) | 28.7 (1.6) | 28.0 (1.4) | 28.5 (2.0) | 28.9 (1.7) |

\*The SVC group has not included for the statistical analysis.

In parallel, a subgroup of participants was investigated and classified by baseline anti-N IgG level (day 0). All participants were divided into two populations, including the seronegative (n=141) and seropositive (n=62) anti-N IgG populations. Participants (n=6) who received a breakthrough infection (defined by anti-N IgG levels) were excluded from the subgroup analysis.

Thus, the number of participants (seronegative: seropositive) were 5:0, 31:16, 34:12, 37:10, and 34:18 for the SVC, SPC, AZC, PFC, and SAC groups, respectively. The demographics and characteristics of the categorized participants are shown in Table 2.

**Table 2.** Demographic and baseline characteristics of the participants classified using seronegative and seropositive anti-N IgG.

| **Population 1: Seronegative anti-N IgG participants (n=141)** |  |  |  |  |  |
| --- | --- | --- | --- | --- | --- |
|  | **SVC\*** | **SPC** | **AZC** | **PFC** | **SAC** |
| **Total number** | 5 | 31 | 34 | 37 | 34 |
| **Sex, Female/total (%)** | 2/5 (40.0) | 14/31 (45.2) | 20/34 (58.8) | 23/37 (62.1) | 11/34 (32.4) |
| **Mean age in year (SD)** | 46.0 (11.2) | 42.6 (13.4) | 50.7 (15.4) | 42.9 (19.4) | 38.9 (13.6) |
| **Interval between doses 2 and 3** |  |  |  |  |  |
| **Mean (SD)** | 367.0 (22.3) | 242.7 (32.1) | 222.9 (43.6) | 184.1 (65.4) | 242.4 (287) |
| **Duration of sample collection** |  |  |  |  |  |
| **(days)** | 14.0 (0.7) | 14.2 (1.1) | 13.7 (1.3) | 14.1 (1.0) | 13.5 (1.0) |
| **Mean (SD) of visit 2** | 27.8 (0.5) | 28.6 (1.9) | 28.1 (1.6) | 28.4 (2.0) | 29.2 (2.0) |
| **Mean (SD) of visit 3** |  |  |  |  |  |
| **Population 2: Seropositive anti-N IgG participants (n=56)** |  |  |  |  |  |
|  | **SVC\*** | **SPC** | **AZC** | **PFC** | **SAC** |
| **Total number** | 0 | 16 | 12 | 10 | 18 |
| **Sex, Female/total (%)** | nd | 5/16 (31.3) | 7/12 (58.3) | 4/10 (40.0) | 12/18 (66.7) |
| **Mean age in year (SD)** | nd | 35.1 (8.1) | 50.9 (12.0) | 41.1 (16.2) | 35.9 (14.8) |
| **Interval between doses 2 and 3** |  |  |  |  |  |
| **Mean (SD)** | nd | 224.4 (34.0) | 209.9 (24.0) | 192.5 (66.2) | 219.8 (32.1) |
| **Duration of sample collection** |  |  |  |  |  |
| **(days)** |  |  |  |  |  |
| **Mean (SD) of visit 2** | nd | 14.1 (0.9) | 14.2 (0.9) | 12.2 (4.7) | 13.8 (0.4) |
| **Mean (SD) of visit 3** | nd | 28.8 (1.2) | 28.0 (0.4) | 25.6 (9.5) | 28.4 (0.8) |

nd – not determined.
\*The SVC group has not included for the statistical analysis.
Six participants who has seroconversion of anti-N IgG after a booster dose were excluded for analysis.

### 3.2 A safety and tolerability profile

Profiles of grades of local and systemic reactions within 7 days after Covovax^™^ vaccination in each primed group had similar incidences (Supplementary Figure S1A–D). A total of 202 of 215 participants experienced a few adverse events. Adverse events were commonly observed as injection site pain (33.7%) rather than myalgia (21.3%), headache (11.8%), and redness (8.9%). In addition, other adverse events were observed to be less than 7% and well tolerated within a few days (Figure 2). One participant in the SVC regimen reported severe injection site pain and headache after the booster dose. None of the participants reported serious adverse events.

**Figure 2.**
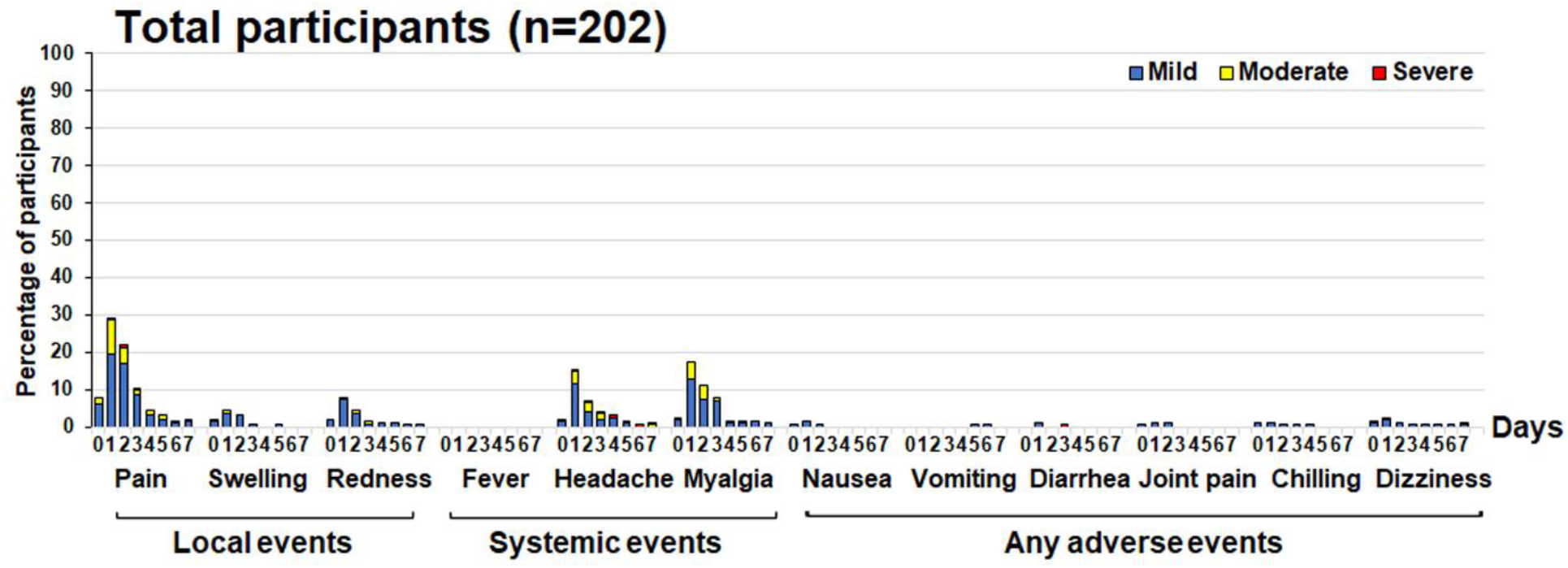
Local, systemic, and adverse events of all participants experienced 7 days after receiving of a protein subunit vaccine.

### 3.3 Anti-RBD Ig/IgG responses of all enrolled participants, negative anti-N IgG participants, and subgroup analysis

Overall, antibody responses were detected in all participants who received the Covovax^™^ vaccine. There were significant differences in anti-RBD Ig GMTs between all groups compared to those receiving SPC at baseline. Before vaccination, the GMTs of the anti-RBD Ig of each of the five groups SVC, SPC, AZC, PFC, and SAC were 17.1, 72.6, 623.5, 1386, and 674.8 U/mL, respectively. Consequently, the GMT of the anti-RBD Ig at 14 days after a booster dose was 18,626, 14,071, 8745, 9539 and 11,286 U/mL for the SVC, SPC, AZC, PFC, and SAC groups, respectively. Subsequently, anti-RBD Ig levels were reduced slightly to 14,410, 9119, 6868, 8564, and 8488 U/mL at 28 days, respectively. The anti-RBD Ig of SPC was significantly higher than AZC, while other groups were comparable at 28 days postvaccination (Figure 3A and Supplementary Table S1). Similar trends were observed with anti-RBD IgG levels (Figure 3B and Supplementary Table S1). However, anti-RBD IgG titers of the SPC group were significantly higher than those of the other three groups at 28 days.

**Figure 3.**
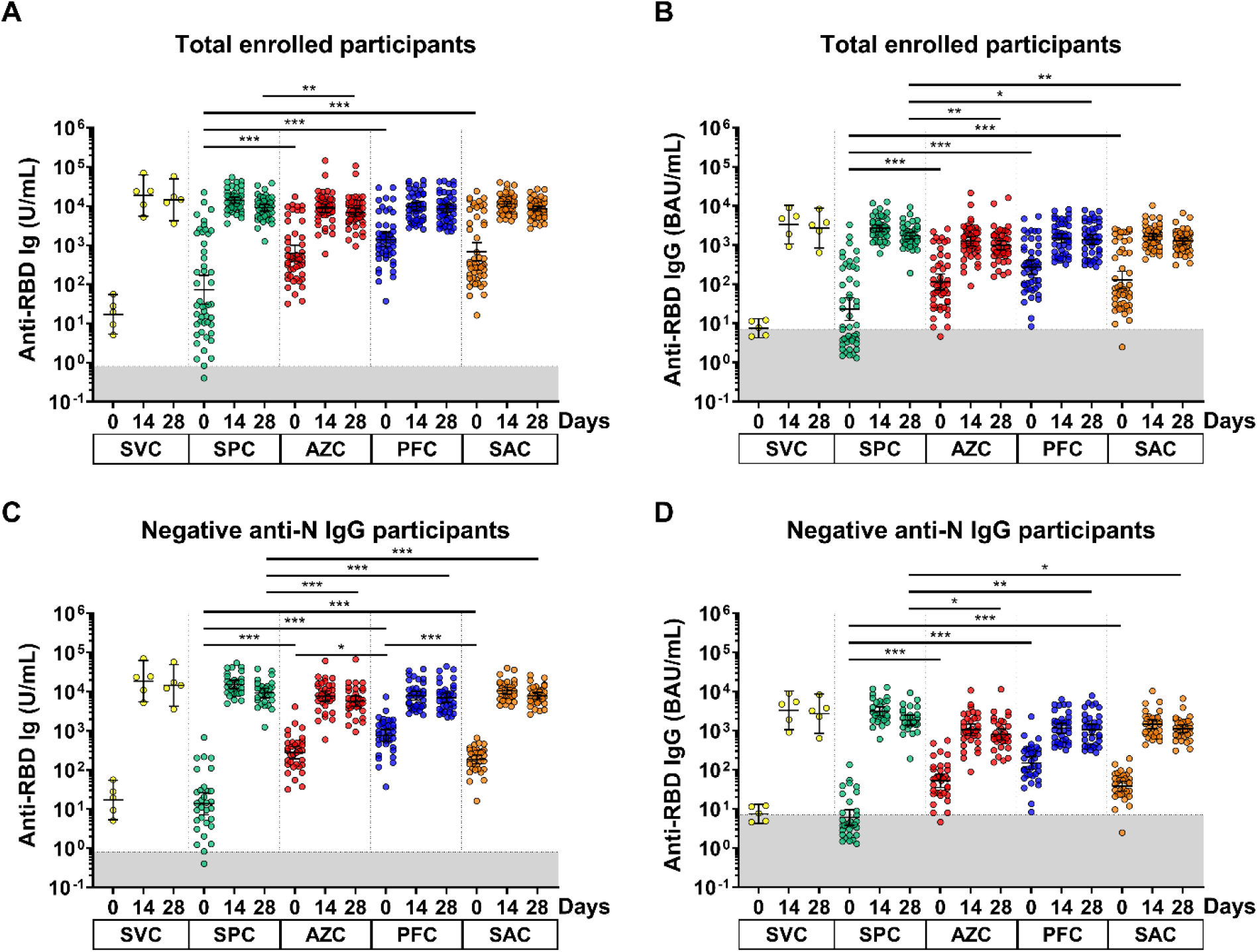
anti-RBD Ig/IgG responses of the enrolled participants. Sera samples of SVC, SPC, AZC, PFC, and SAC groups were monitored at 0, 14 and 28 days. (A) anti-RBD Ig (U/mL) and (B) anti-RBD IgG (BAU/mL) of the total enrolled participants. (C) anti-RBD Ig (U/mL) and (D) anti-RBD IgG (BAU/mL) of the seronegative anti-N IgG participants. The lines represent GMTs (95% confidence intervals). The gray area indicates the seronegativity of the anti-RBD Ig (<0.8 U/mL) or anti-RBG IgG (<7.1 BAU/mL). *p* <0.05 (*), *p* <0.01 (**), *p* <0.001 (***).

Based on the baseline anti-RBD Ig/IgG titers (Figure 3A-B), participants could be stratified into two populations. This population could be further classified based on anti-N IgG results. Overall, 141/197 (71.6%) individuals were seronegative for anti-N IgG (< 1.40 S/C). Here, we further analyzed the anti-RBD Ig/IgG titers of seronegative participants for anti-N IgG, as indicated in Figure 3C,D. The baseline GMTs of anti-RBD Ig were 17.1, 13.7, 276.2, 759.6, and 186.7 U/mL for the respective groups. At baseline, the GMT of anti-RBD Ig was significantly lower in the SPC group compared to others. At 14 days, the GMTs of anti-RBD Ig were 18626, 15149, 7627, 7846, and 10494 U/mL, respectively and consequently, the anti-RBD IgG levels were slightly reduced to 14410, 9270, 5735, 7032, and 7865 U/mL at 28 days, respectively. Furthermore, comparable anti-RBD IgG levels were determined for all groups except for those receiving SPC, whose titers showed a significant difference (Figure 3C and Supplementary Table S1). Moreover, a similar response was observed anti-RBD IgG levels (Figure 3C and Supplementary Table S1).

In total, 192 individuals receiving SPC, AZC, PFC, and SAC vaccines were classified by subgroup analysis according to the baseline anti-N IgG levels. Five individuals receiving SVC were not included in the calculation. The anti-RBD Ig levels between seronegative and seropositive anti-N IgG populations was compared. At baseline, the anti-RBD Ig of the seropositive population in all groups was significantly higher than that of the seronegative population. After a booster dose, the anti-RBD Ig of the SPC and SAC regimens showed a significant difference between populations (Supplementary Figures S2A,D). In contrast, there was no significant difference in AZC and PFC regimens (Supplementary Figure S2B,C).

### 3.4 Neutralizing activity against wild type and omicron BA.2 using sVNT

At baseline, the number of seropositive neutralizing activities against the wild type SARS-CoV-2 strain following SVC, SPC, AZC, PFC, and SAC regimes were 0/10, 0/10, 7/10, 8/10, and 2/10, respectively (Figure 4A and Supplementary Table S1). Nonetheless, some seropositive neutralizing activities against the BA.2 omicron variant of the respective groups were also detected in 0/10, 0/10, 1,10, 7/10 and 0/10, respectively (Figure 4B and Supplementary Table S1).

**Figure 4.**
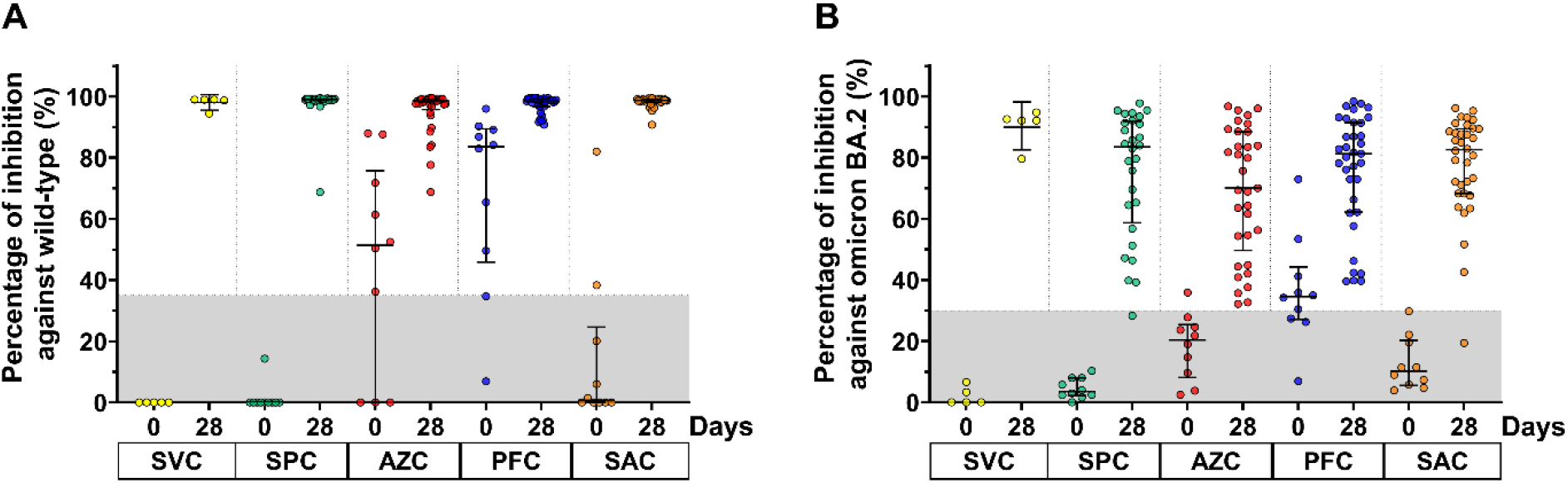
Neutralizing activity of participants against the wild type SARS-CoV-2 virus (A) and BA.2 omicron variant (B). Lines represent the median (interquartile range [IQR]). The gray area indicates the seronegativity of neutralizing activity of the wild type (<35%) and BA.2 omicron variant (<30%).

At day 28 after a booster dose, all boosted participants showed restored neutralizing activity against the wild type by more than 95% inhibition, reaching the upper limit of an assay (Figure 4A). The neutralizing activity against the omicron BA.2 variant in participants achieved marked inhibitory activity as indicated by the 92.1%, 83.5%, 70.1%, 81.3%, and 82.6% values corresponding to SVC, SPC, AZC, PFC, and SAC, respectively. Only two participants showed no seroconversion of neutralizing activity against omicron BA.2. Furthermore, the percentage inhibition against BA.2 omicron variant in all regimens was comparable (Figure 4B).

### 3.5 Total IFN-γ release induced by the Ag3 QFN assay

The T-cell response was assessed by measuring total IFN-γ release in whole blood samples of participants who received a booster dose. At baseline, the remaining IFN-γ level of IFN-after the second dose was less than 0.2 IU/mL for all participants. The results showed that the IFN-γ level after vaccination of all participants was significantly increased at 14 days and slightly reduced at the 28-day follow-up except for those receiving the SAC regime. The group comparison indicated that the IFN-γ level was comparable at 14 and 28 days (Figure 5). The elevated response at 14 days was 5/5 (100%), 19/21 (90.4%), 18/21 (85.7%), 17/24 (70.8%), and 18/22 (81.8%) for the respective groups.

**Figure 5.**
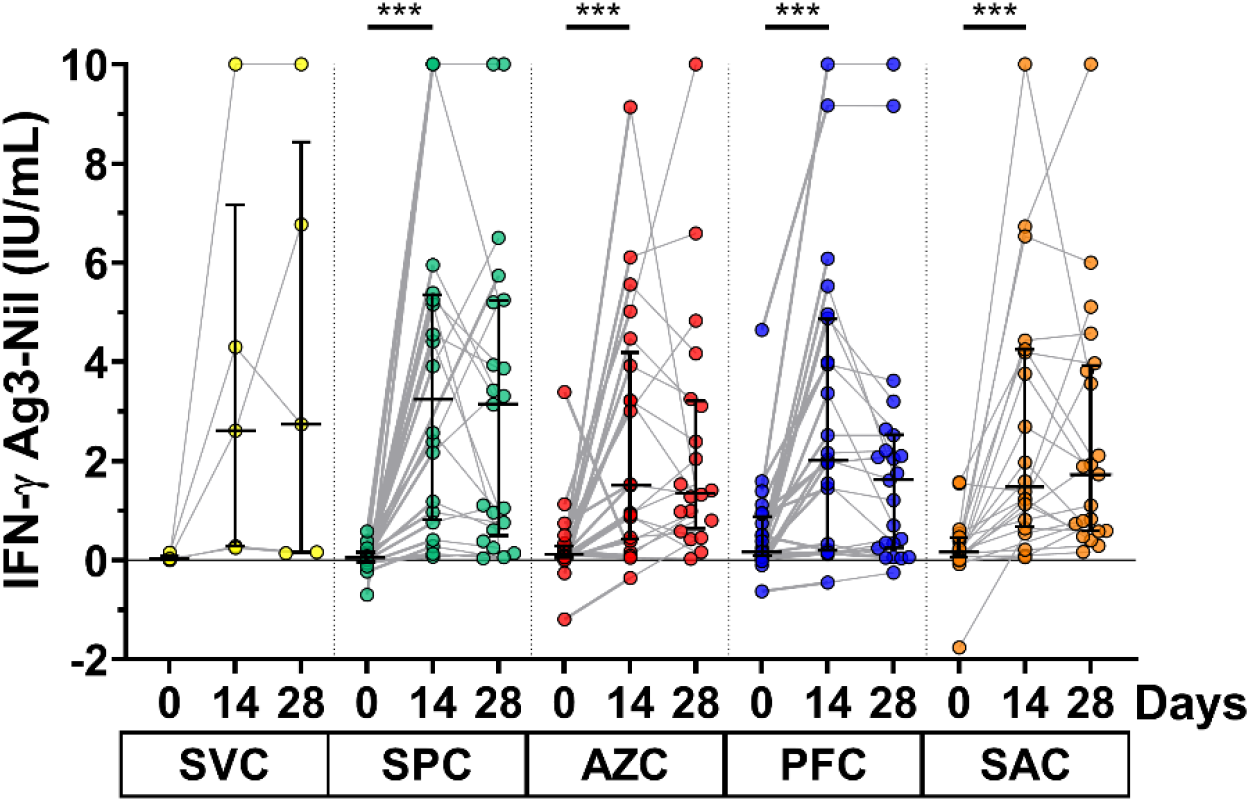
Interferon-gamma release assay. Whole blood samples from the participants were heparinized and then transferred to the Ag3-QFN blood collection tube for 21 hours. The IFN-γ release was monitored by ELISA. Lines represent the median (IQR). *p* <0.05 (*), *p* <0.01 (**), *p* <0.001 (***).

## 4. Discussion

Herein, we report the first study evaluating responses to the Covovax^™^ protein subunit vaccine as a booster dose in individuals who had received two-dose vaccination (priming) with either homologous inactivated or heterologous inactivated/viral vector vaccine regimens. The reactogenicity and immunogenicity of the protein subunit vaccine. The recruitment criteria were no history of COVID-19 infection. The enrolled participants included a seropositive anti-N IgG population of approximately 30% during the study period (May 2022 to July 2022). Individuals with a positive anti-N IgG may have experienced an infection leading to asymptomatic or mild disease. The adverse events record revealed common effects were injection site pain, myalgia, headache, and redness at the injection site. A few incidences of adverse effects were observed but were assessed as mild-to-moderate in severity. The adverse events reported herein were similar to those of pain, myalgia, and headache described in a previous Novavax study from Australia (Keech et al., 2020, Mallory et al., 2022) and the United States (Mallory et al., 2022). Furthermore, our findings demonstrated that the Covovax^™^ vaccine elicited fewer adverse events compared to the viral vector or the mRNA-COVID-19 vaccine (Assawakosri et al., 2022a, Fiolet et al., 2022, Munro et al., 2021). Furthermore, the adverse effects following vaccination differed based on the booster vaccine administered. There was no correlation with the history of the previous vaccine.

Before receiving booster doses, the anti-RBD Ig/IgG titer in immunized participants was notably lower. The anti-RBD Ig and IgG responses after boost indicated that the levels of binding antibodies were significantly increased regardless of the primary series vaccine regimen received. Our study found that the Covovax^™^ vaccine was suitable for use as a booster dose. Consistent with a previous study (Munro et al., 2021), the Novavax subunit protein vaccine could elicit a high antispike protein IgG titer in individuals who had received two doses of AZD1222 or BNT162b2.

Compared to previous studies evaluating three doses of vaccination in healthy adults, we observed that using the Covovax^™^ vaccine in the primary series of the inactivated vaccine group achieved higher antibody levels than three doses of the inactivated vaccine (Ai et al., 2022, Assawakosri et al., 2022b). Furthermore, a heterologous booster dose including Covovax^™^ in the primary series of viral vector vaccine (AZD1222) achieved a robust immune response compared to three doses of AZD1222 (Assawakosri et al., 2022a). However, a heterologous booster dose with Covovax^™^ in individuals receiving a primary series of BNT162b2 showed a lower immune response compared to three doses of BNT62b2 (Wanlapakorn et al., 2022a). Notably, multiple factors, such as sex, age, and duration after the primary series, might have interfered with the analysis of the booster effect.

Consistent with the anti-RBD Ig and IgG responses observed in this study, the booster dose elicited a high immune response and neutralizing activity against wild type and omicron BA.2 variants of SARS-CoV-2. However, neutralizing antibodies against the omicron variant were slightly lower than that toward the prototype variant (Bhiman et al., 2022). Several mutations in the RBD could alter the binding affinity of antibodies produced from wild type vaccines (Cao et al., 2022). The present study observed IFN-γ releasing after 14 days among all regimens. In agreement with previous studies, the protein subunit vaccine and Matrix-M1 adjuvant could induce high levels of cellular immunity (Keech et al., 2020, Tian et al., 2021). Besides the humoral response, the Covovax^™^ vaccine could elicit a robust cellular response.

The Com-COV trial in the United Kingdom examined sera for anti-N IgG levels at baseline and used seropositivity to define SARS-CoV-2 infection (Lui et al., 2021). Nearly one-third of the participants were suspected of exposure to SARS-CoV-2 infection prior to enrollment. These individuals were associated with asymptomatic infection or mild/transient symptoms, indicating an underestimate of SARS-CoV-2 infection (Tande et al., 2022). The subgroup analysis may suggest that both seronegative and seropositive anti-N IgG levels enhanced the anti-RBD Ig titer as seen in the AZC and PFC groups. In contrast, differences in the SPC and SVC vaccinated groups were statistically significant. Furthermore, participants in this study had experienced SARS-CoV-2 infection within a few weeks of sampling, despite having received booster vaccination.

Our study was subject to certain limitations. First, the SCV group contained a lower number of participants due to Thailand’s enacting of a massive CoronaVac vaccination campaign since April 2021, which has diminished further after mRNA vaccines were implemented in September 2021. This population had received a third dose of another vaccine to address the third and subsequent waves of the SARS-CoV-2 endemic. Furthermore, the surrogate virus neutralization test against the wild type reached the upper limit of >95% inhibition (Kanokudom et al., 2022). The pseudovirus reduction neutralization test will be used in future to overcome these limitations. Additionally, we were unable to obtain the results of COVID-19 testing of the seropositive anti-N IgG population to establish their history of SARS-CoV-2 infection.

## 5. Conclusions

The protein subunit vaccine as a booster dose is safe and elicits a high level of binding and neutralizing antibodies against SARS-CoV-2, as well as a good cellular response. Thus, the Covovax^™^ vaccine can be recommended for use as a booster in individuals receiving variable primary vaccine regimens. Our data indicate the Covovax^™^ vaccine will be beneficial and convenient as a heterologous booster dose and will be of value for public health vaccine implementation guidelines.

## Data Availability

All data produced in the present study are available upon reasonable request to the authors

## Acknowledgments

We would like to thank all Center of Excellence in Clinical Virology personnel and all participants for contributing to and supporting this project. This research was financially supported by the Health Systems Research Institute (HSRI), National Research Council of Thailand (NRCT), the Center of Excellence in Clinical Virology, Chulalongkorn University, and King Chulalongkorn Memorial Hospital, and partially supported by the Second Century Fund (C2F) of Sitthichai Kanokudom, Chulalongkorn University.

## Author Contributions

Conceptualization, S.K. (Sitthichai Kanokudom), P.N., S.H. and Y.P.; data curation, S.K. (Sitthichai Kanokudom), R.Y., N.S. (Nungruthai Suntronwong), S.A., A.K., W.T., J.A., W.W., D.S., T.T. (Thaksaporn Thatsanatorn), N.S. (Natthinee Sudhinaraset) and N.W.; formal analysis, S.K. (Sitthichai Kanokudom); methodology, S.K. (Sitthichai Kanokudom), J.C., R.A., N.K., P.V., S.K. (Sirapa Klinfueng) and T.T (Thanunrat Thongmee); project administration, Y.P.; writing—original draft, S.K. (Sitthichai Kanokudom); writing— review and editing, S.K. (Sitthichai Kanokudom), S.H. and Y.P. All authors have read and agreed to the published version of the manuscript.

## Institutional Review Board Statement

The study protocol was approved by the Institutional Review Board (IRB), Faculty of Medicine, Chulalongkorn University (IRB number 871/64).

## Informed Consent Statement

Informed consent was obtained before participant enrollment. The study was conducted according to the Declaration of Helsinki and the Good Clinical Practice Guidelines (ICH-GCP) principles.

## Data Availability Statement

The datasets generated and analyzed during the current study are available from the corresponding author upon reasonable request.

## Supplementary materials

**Table S1** Descriptive analysis of laboratory assessments

**Figure S1**. Local, systemic, and adverse events following different prime vaccination regimens 7 days after receiving a booster dose. (A) SPC, (B) AZC, (C) PFC, and (D) SAC.

**Figure S2**. Comparison of individual anti-RBD Ig values stratified by anti-N IgG titers. Participants were classified by seronegative and seropositive anti-N IgG (Cut-off ≥ 1.4 S/C) before a booster dose. (A) SPC, (B) AZC, (C) PFC, (D) SAC regimens. The lines represent GMTs (95% confidence interval [CI]). The gray area indicates the seronegativity of the anti-RBD Ig (<0.8 U/mL). *p* <0.05 (*), *p* <0.001 (***).

