## Supplementary material for "Safety and immunogenicity of a third dose of COVID-19 protein subunit vaccine (Covovax™) after homologous and heterologous two-dose regimens": Supplementary Materials.docx

**Supplementary materials
Table S1** Descriptive analysis of laboratory assessments

|  | SVC | SPC | AZC | PFC | SAC |
| --- | --- | --- | --- | --- | --- |
| Humoral responses  Anti RBD Ig (U/mL) of all enrolled participants | | | | | |
| Visit 1 (Day 0), n  GMT (95% CI)  GMR (95% CI) | 5  17.1 (5.3-54.7)  nd | 48  72.6 (31.1-169.1)  ref | 49  623.5 (388.2-1002)  0.10 (0.03-0.31)* | 49  1386 (915.4-2099)  0.08 (0.02-0.24)* | 52  674.8 (394.1-1155)  0.08 (0.02-0.24)* |
| Visit 2 (Day 14), n  GMT (95% CI)  GMR (95% CI) | 5  18626 (5544-62571)  nd | 47  14071 (11748-16854)  ref | 49  8745 (6766-11302)  1.91 (1.27-2.89)* | 47  9539 (7500-12134)  1.87 (1.21-2.90)* | 50  11286 (9693-13141)  1.50 (0.99-2.27) |
| Visit 3 (Day 28), n  GMT (95% CI)  GMR (95% CI) | 5  14410 (4226-49134)  nd | 45  9119 (7466-11139)  ref | 48 6868 (5337-8838)  1.65 (1.09-2.51)* | 47  8564 (6667-11002)  1.43 (0.92-2.22) | 51  8488 (7300-9871)  1.35 (0.89-2.04) |
| Anti RBD IgG (BAU/mL) of all enrolled participants | | | | | |
| Visit 1 (Day 0), n  GMT (95% CI)  GMR (95% CI) | 5  7.5 (4.3-13.2)  nd | 48  23.0 (11.8-44.8)  ref | 49  113.9 (71.5-181.5)  0.19 (0.07-0.52)* | 49  275.0 (179.9-420.5)  0.14 (0.05-0.39)* | 52  128.0 (76.6-214.0)  0.19 (0.08-0.48)* |
| Visit 2 (Day 14), n  GMT (95% CI)  GMR (95% CI) | 5  3325 (1069-10341)  nd | 47  2707 (2229-3287)  ref | 49  1249 (947.5-1648)  2.35 (1.48-3.73)* | 47  1483 (1134-1939)  2.31 (1.43-3.72)* | 50  1642 (1364-1976)  2.02 (1.30-3.15)* |
| Visit 3 (Day 28), n  GMT (95% CI)  GMR (95% CI) | 5  2720 (848.5-8718)  nd | 45  1727 (1418-2103)  ref | 48  989.0 (753.5-1298)  1.88 (1.19-2.96)* | 47  1377 (1036-1830)  1.63 (1.02-2.59)* | 51  1270 (1068-1511)  1.69 (1.10-2.60)* |
| Anti RBD Ig (U/mL) of a seronegative anti-N IgG participant | | | | | |
| Visit 1 (Day 0), n  GMT (95% CI)  GMR (95% CI) | 5  17.1 (5.3-54.7)  nd | 31  13.7 (7.2-26.1)  ref | 34  276.2 (191.9-397.5)  0.04 (0.02-0.08)* | 37  759.6 (541.3-1066)  0.02 (0.01-0.04)* | 34  186.7 (143.5-243.0)  0.08 (0.04-0.17)* |
| Visit 2 (Day 14), n  GMT (95% CI)  GMR (95% CI) | 5  18626 (5544-62571)  nd | 30  15149 (11972-19170)  ref | 34  7627 (5564-10454)  3.36 (1.78-6.34)* | 36  7846 (6063-10152)  3.90 (1.87-8.13)* | 32  10494 (8522-12922)  2.29 (1.24-4.19)* |
| Visit 3 (Day 28), n  GMT (95% CI)  GMR (95% CI) | 5  14410 (4226-49134)  nd | 28  9270 (6961-12343)  ref | 33  5735 (4237-7764)  2.47 (1.23-4.93)* | 36  7032 (5342-9258)  2.54 (1.17-5.48)* | 33  7865 (6444-9600)  1.87 (1.01-3.47)* |
| Anti RBD IgG (BAU/mL) of a seronegative anti-N IgG participant | | | | | |
| Visit 1 (Day 0), n  GMT (95% CI)  GMR (95% CI) | 5  7.5 (4.3-13.2)  nd | 31  6.1 (3.9-9.6)  ref | 34 52.6 (35.4-78.1)  0.11 (0.06-0.22)* | 37  149.4 (103.3-216.3)  0.06 (0.03-0.13)* | 34  38.3 (28.7-51.2)  0.17 (0.09-0.33)* |
| Visit 2 (Day 14), n  GMT (95% CI)  GMR (95% CI) | 5  3325 (1069-10341)  nd | 30  3140 (2421-4073)  ref | 34  1054 (749.1-1482)  5.21 (2.75-9.84)* | 36 1159 (868.3-1548)  6.24 (2.96-13.15)* | 32  1463 (1125-1903)  3.48 (1.87-6.43)* |
| Visit 3 (Day 28), n  GMT (95% CI)  GMR (95% CI) | 5  2720 (848.5-8718)  nd | 28  1876 (1405-2504)  ref | 33  799.4 (577.0-1108)  3.45 (1.77-6.75)* | 36  1078 (785.9-1478)  3.52 (1.65-7.55)* | 33  1122 (891.2-1413)  2.60 (1.43-4.74)* |
| Anti RBD Ig (U/mL) of a seropositive anti-N IgG participant | | | | | |
| Visit 1 (Day 0), n  GMT (95% CI) | 0  nd | 16  1934 (753.5-4963) | 12  6582 (4208-10296) | 10  9758 (5539-17191) | 18  7643 (5287-11048) |
| Visit 2 (Day 14), n  GMT (95% CI) | 0  nd | 16  11690 (8709-15691) | 12  9799 (7285-13180) | 9  17566 (10496-29399) | 18  12845 (10338-15960) |
| Visit 3 (Day 28), n  GMT (95% CI) | 0  nd | 16  8511 (6401-11316) | 12  8183 (6011-11141) | 9  14834 (8990-24474) | 18  9763 (7735-12322) |
| Percentage of inhibition against wild type (%) | | | | | |
| Visit 1 (Day 0), n  Med (IQR) | 5  0.0 (0.0-0.0) | 10  0.0 (0.0-0.0) | 10  51.5 (0.0-75.8) | 10  83.6 (45.9-89.5) | 10  0.7 (0.0-24.7) |
| Visit 3 (Day 28), n  Med (IQR) | 5  99.0 (96.6-99.0) | 28  99.1 (98.8-99.2) | 33  98.6 (95.7-99.1) | 35  98.5 (96.7-99.2) | 34  98.8 (98.1-99.0) |
| Percentage of inhibition against omicron BA.2 (%) | | | | | |
| Visit 1 (Day 0), n  Med (IQR) | 5  0.0 (0.0-5. 0) | 10  3.5 (2.2-8.0) | 10  20.4 (8.2-25.4) | 10  34.6 (27.1-44.3) | 10  10.2 (5.5-20.2) |
| Visit 3 (Day 28), n  Med (IQR) | 5  92.1 (85.9-93.7) | 28  83.5 (58.7-91.9) | 33  70.1 (49.7-88.6) | 35  81.3 (62.2-91.5) | 34  82.6 (68.3-89.5) |
| Cellular response  IFN- γ Ag3-nil (IU/mL) | | | | | |
| Visit 1 (Day 0), n  Med (IQR) | 5  0.0 (0.0-0.1) | 21  0.0 (0-0.1) | 21  0.1 (0.0-0.3) | 24  0.2 (0.1-0.9) | 22  0.1 (0.0-0.5) |
| Visit 2 (Day 14), n  Med (IQR) | 5  2.6 (0.3-7.2) | 20  3.2 (0.8-5.4) | 21  1.5 (0.4-4.2) | 23  2.0 (0.2-4.9) | 20  1.5 (0.7-4.2) |
| Visit 3 (Day 28), n  Med (IQR) | 5  2.7 (0.2-8.4) | 21  3.1 (0.5-5.2) | 20  1.4 (0.6-3.2) | 23  1.6 (0.3-2.5) | 21  1.7 (0.6-3.9) |

GMR refers to the geometric mean ratio. * The mean difference is significant at the 0.05 level. nd – not determined. ref – referent group was used as comparative group.

**Supplementary Figures**

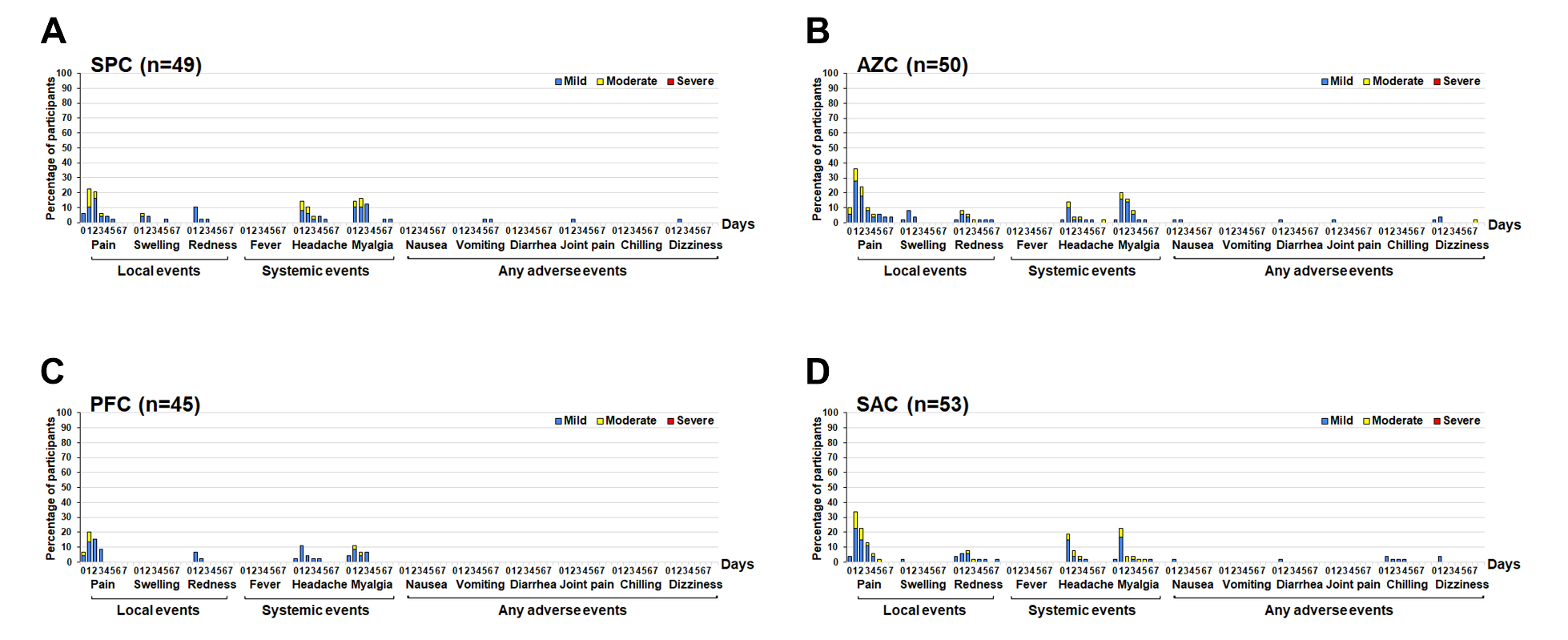

**Figure S1.** Local, systemic, and adverse events following different prime vaccination regimens 7 days after receiving a booster dose. (A) SPC, (B) AZC, (C) PFC, and (D) SAC.

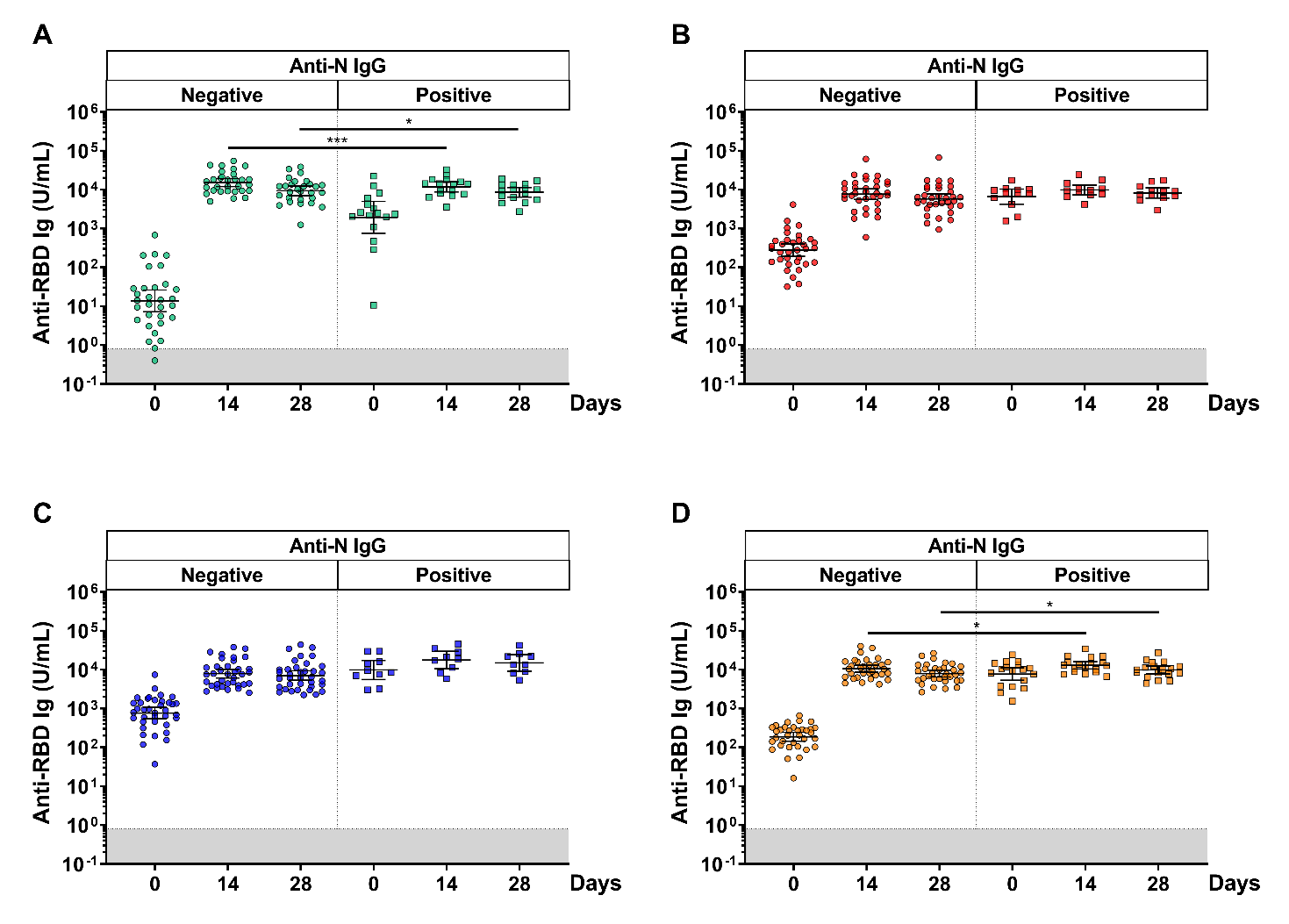

**Figure S2.** Comparison of individual anti-RBD Ig values stratified by anti-N IgG titers. Participants were classified by seronegative and seropositive anti-N IgG (Cut-off ≥ 1.4 S/C) before a booster dose. (A) SPC, (B) AZC, (C) PFC, (D) SAC regimens. The lines represent GMTs (95% confidence interval [CI]). The gray area indicates the seronegativity of the anti-RBD Ig (<0.8 U/mL). *p* <0.05 (*), *p* <0.001 (***).
